# Persistent Atrial Myopathy Despite Ventricular Recovery: Prognostic Significance of Discordant LV–LA Strain Patterns in HFrEF

**DOI:** 10.64898/2026.04.22.26351480

**Authors:** Jiesuck Park, Nan Young Bae, Jaehyun Lim, Soongu Kwak, MinJung Bak, Hong-Mi Choi, Jun-Bean Park, Yeonyee E. Yoon, Seung-Pyo Lee, Yong-Jin Kim, Goo-Yeong Cho, Hyung-Kwan Kim, In-Chang Hwang

**Affiliations:** Department of Cardiology, Cardiovascular Center, Seoul National University Bundang Hospital, Republic of Korea; Department of Internal Medicine, Seoul National University College of Medicine, Republic of Korea; Healthcare System Gangnam Center, Seoul National University Hospital, Republic of Korea; Cardiovascular Center and Department of Internal Medicine, Seoul National University Hospital, Republic of Korea

**Keywords:** angiotensin receptor–neprilysin inhibitor, heart failure, left atrial reservoir strain, left ventricular global longitudinal strain, reverse remodelling, prognosis

## Abstract

**Aims:** Assessment of treatment response in HFrEF has largely relied on left ventricular (LV)-centric parameters, yet the left atrium (LA) modulates LV filling and reflects the cumulative hemodynamic burden. Whether discordant recovery between LV and LA function carries distinct prognostic implications in patients treated with ARNI-based therapy remains unknown.

**Methods and results:** From the multicentre STRATS-HF-ARNI registry, 1,182 patients with HFrEF who underwent serial echocardiography at baseline and one-year follow-up were included. Patients were classified according to the change in LVGLS and LASr at one year: Group A, concordant recovery (57.4%); Group B, discordant atrial non-recovery (11.2%); Group C, discordant ventricular non-recovery (15.6%); and Group D, concordant non-recovery (16.0%). Outcomes included all-cause mortality, cardiovascular mortality, and HF hospitalization. Despite achieving LV functional improvement, Group B exhibited persistent LASr deterioration, accompanied by less favourable hemodynamic trajectories than Group A. On multivariable Cox regression, Group B had significantly higher risks of all-cause mortality (adjusted hazard ratio [aHR] 3.53, 95% confidence interval [CI] 1.60–7.79) and cardiovascular mortality (aHR 5.68, 95% CI 1.91–16.92), comparable to Group D. Group C demonstrated higher HF hospitalization risk (aHR 2.25, 95% CI 1.31–3.86). The adverse prognostic impact of discordant atrial non-recovery was consistently observed across subgroups stratified by baseline LVGLS and LASr levels.

**Conclusion:** In HFrEF patients treated with ARNI-based therapy, persistent LA dysfunction despite LV functional improvement identifies a high-risk phenotype comparable to concordant non-recovery. These findings suggest that concurrent assessment of LV and LA strain may provide incremental prognostic value beyond LV-centric metrics alone.

## 1. INTRODUCTION

Heart failure (HF) is characterized by progressive structural and functional remodelling of both the left ventricle (LV) and left atrium (LA), reflecting the hemodynamic stress and neurohormonal activation. While contemporary guideline-directed medical therapy—including angiotensin receptor–neprilysin inhibitors (ARNIs)—has been shown to promote reverse remodelling and improve clinical outcomes,^1,2^ the assessment of treatment response has largely relied on LV-centric parameters, particularly left ventricular ejection fraction (LVEF) and LV global longitudinal strain (LVGLS).^3,4^ This ventricular-centric paradigm, however, may incompletely capture the full scope of myocardial and hemodynamic recovery in HF.

The LA plays a central role in modulating LV filling and encodes the cumulative hemodynamic burden of elevated filling pressures. LA reservoir strain (LASr), a sensitive speckle-tracking–derived measure of LA function, integrates the effects of diastolic dysfunction and atrial remodelling and has emerged as a robust and independent prognostic marker across diverse HF populations.^5,6^ In contrast to LV function, which demonstrates relatively prompt improvement with ARNI-based therapy, LA dysfunction often reflects more advanced or chronic myocardial disease with less consistent reversibility. Accordingly, discordance between LV and LA functional recovery may reveal residual disease activity not captured by conventional LV-focused metrics, with distinct implications for risk stratification.^7^

Recent studies have underscored the prognostic importance of strain imaging in HF; however, the longitudinal interplay between LV and LA functional recovery following ARNI therapy remains insufficiently characterized.^8,9^ In particular, it is unclear whether improvement in LV systolic function alone adequately reflects true myocardial recovery, or whether persistent LA dysfunction identifies a high-risk phenotype despite apparent ventricular improvement. This concept of “discordant recovery”—wherein LVGLS and LASr diverge in their trajectories under treatment—has not been systematically evaluated in a large, real-world cohort of patients with heart failure with reduced ejection fraction (HFrEF).

In the present study, we classified patients with HFrEF treated with ARNI-based therapy according to recovery patterns in LVGLS and LASr at one-year follow-up, using data from a multi-centre real-world HF registry. We further evaluated the prognostic implications of each recovery phenotype for subsequent clinical outcomes.

## 2. METHODS

### 2.1 Study Population

Clinical data were retrospectively analysed from the STrain for Risk Assessment and Therapeutic Strategies in patients with Heart Failure treated with Angiotensin Receptor-Neprilysin Inhibitor (STRATS-HF-ARNI) registry. The STRATS-HF-ARNI registry enrolled 2,757 consecutive patients diagnosed with HFrEF who initiated ARNI-based therapy at two tertiary medical centres in Korea (Seoul National University Hospital and Seoul National University Bundang Hospital) between 2017 and 2022. The STRATS-HF-ARNI study was registered with the Clinical Research Information Service of the Ministry of Health and Welfare of the Republic of Korea (registration number: KCT0008098), and details of the registry design and patient characteristics have been described previously.^10,11^

For the present analysis, patients were eligible if they had a confirmed diagnosis of HFrEF at baseline and underwent serial transthoracic echocardiography (TTE) at both baseline and one-year follow-up. The following exclusion criteria were applied: absence of a baseline TTE (n=336), absence of a one-year follow-up TTE (n=1,180), missing LVGLS or LASr measurements at either timepoint (n=28), and absence of clinical follow-up data (n=31). After applying these criteria, a final cohort of 1,182 patients was included in the analysis (**Figure 1**). The study protocol was approved by the Institutional Review Boards of Seoul National University Hospital (J-2212-034-1383) and Seoul National University Bundang Hospital (B-2005-615-108), with a waiver of informed consent granted due to the retrospective study design.

**Figure 1.**
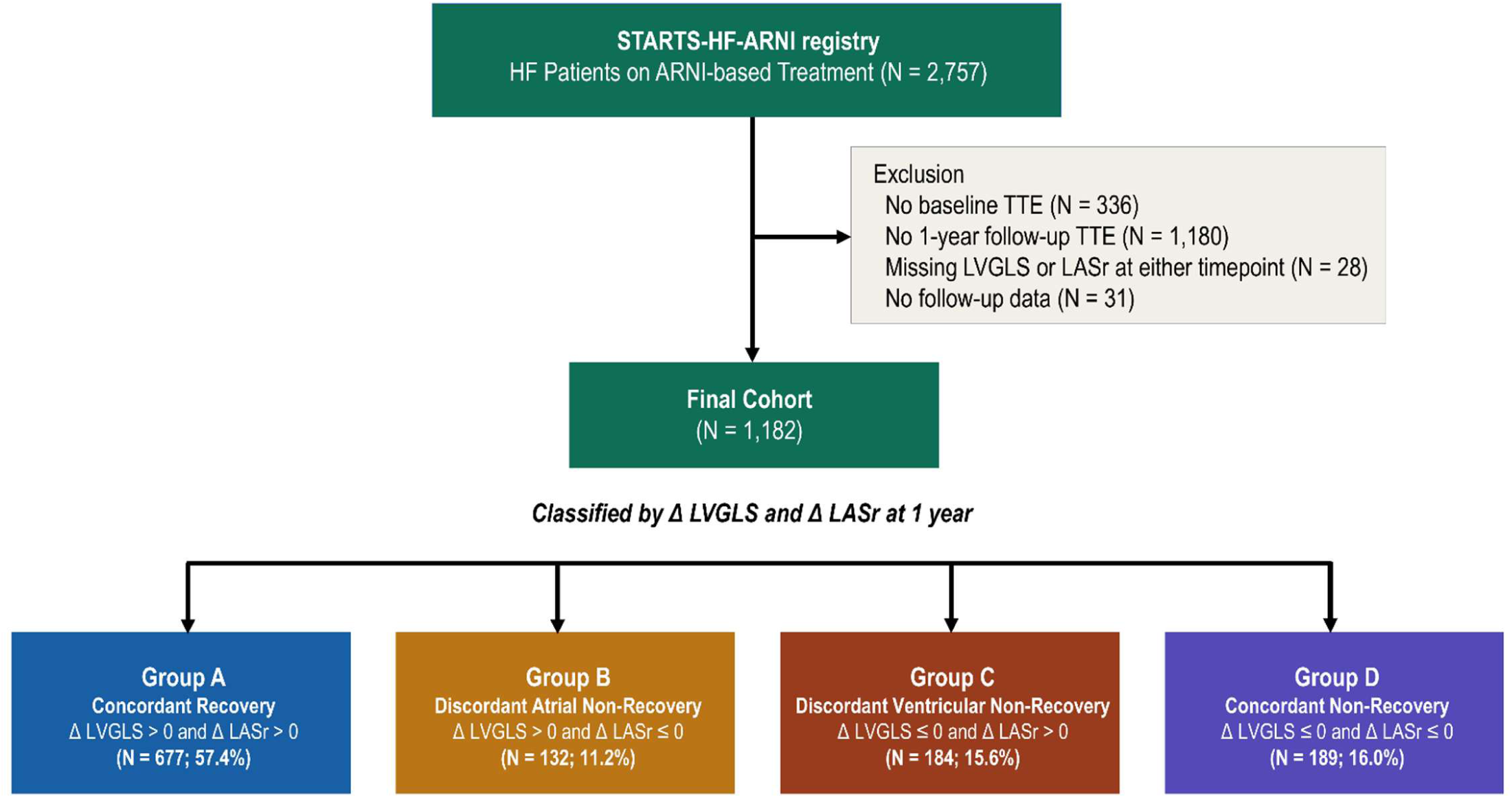
Study Flow. Of 2,757 consecutive patients with HFrEF on ARNI-based treatment in the STRATS-HF-ARNI registry, 1,182 patients with available LVGLS and LASr measurements at both baseline and one-year follow-up were included in the final analysis. Patients were classified into four mutually exclusive strain recovery phenotypes according to the direction of change in LVGLS and LASr at one year. ARNI, angiotensin receptor–neprilysin inhibitor; HFrEF, heart failure with reduced ejection fraction; LASr, left atrial reservoir strain; LVGLS, left ventricular global longitudinal strain; TTE, transthoracic echocardiography.

### 2.2 Echocardiographic Evaluation

All echocardiographic examinations were performed by trained echocardiographers or cardiologists and interpreted by board-certified cardiologists specialized in echocardiography, following current guidelines, as part of routine clinical care.^12^ Standard TTE was performed at baseline and at the one-year follow-up visit.

Two-dimensional speckle-tracking echocardiography was performed to quantify myocardial deformation using TomTec software (Image Arena 4.6, Munich, Germany).^13^ LVGLS was derived as the average peak systolic longitudinal strain from all LV myocardial segments, obtained from the apical four-chamber, two-chamber, and apical long-axis views. LASr was defined as the peak positive longitudinal strain of the LA wall during ventricular systole—corresponding to LA reservoir function—and was assessed from the apical four-chamber and two-chamber views. LA contractile strain (LASct) was defined as the change in LA longitudinal strain from the onset of atrial contraction to end-diastole, reflecting LA booster-pump function. These strain parameters were reported as absolute values, with higher values indicating greater myocardial deformation and better chamber function. Serial changes were quantified as ΔLVGLS, ΔLASr and ΔLASct (one-year value minus baseline value), with positive values reflecting functional improvement in the respective chamber.

### 2.3 Classification of Strain Recovery Phenotypes

Patients were classified into four mutually exclusive strain recovery phenotypes according to the direction of change in both LVGLS and LASr at one-year follow-up: Group A, concordant recovery (ΔLVGLS >0 and ΔLASr >0); Group B, discordant atrial non-recovery (ΔLVGLS >0 and ΔLASr ≤0); Group C, discordant ventricular non-recovery (ΔLVGLS ≤0 and ΔLASr >0); and Group D, concordant non-recovery (ΔLVGLS ≤0 and ΔLASr ≤0). Group A was designated as the reference group for all comparative analyses.

### 2.4 Clinical Outcomes

The primary outcome was all-cause mortality. Secondary outcomes included cardiovascular mortality and HF hospitalization. Clinical outcomes were ascertained from the date of one-year echocardiographic follow-up through the end of the observation period via dedicated review of the electronic medical records.

### 2.5 Statistical Analysis

Clinical data are presented as median with interquartile range (IQR) for continuous variables and as frequency with percentage for categorical variables. Differences in baseline characteristics across the four groups were assessed using the Kruskal-Wallis test for continuous variables and the chi-square test for categorical variables. Within-group changes in echocardiographic parameters, including LVGLS and LASr, between baseline and one-year follow-up were assessed using the Wilcoxon signed-rank test. Between-group differences in the magnitude of change (i.e., ΔLVGLS and ΔLASr) were evaluated using the Kruskal-Wallis test.

Cumulative event rates were estimated using the Kaplan-Meier method and compared across groups using the log-rank test. The association between strain recovery phenotype and clinical outcomes was evaluated using Cox proportional hazards regression, with Group A as the reference. Multivariable models were adjusted for age, sex, baseline LVEF, N-terminal pro–B-type natriuretic peptide (NT-proBNP), ischemic cardiomyopathy, chronic kidney disease, and concomitant sodium–glucose cotransporter 2 (SGLT2) inhibitor use, and results are expressed as adjusted hazard ratios (aHR). The prognostic association of LVGLS and LASr changes as continuous variables with clinical outcomes was additionally evaluated using Cox regression, with results reported per 5% decrease in each strain parameter.

Subgroup analyses were performed to examine the association between strain recovery phenotype and clinical outcomes according to baseline LVGLS and LASr level, each dichotomized at the median value. As a sensitivity analysis, a volume-based recovery classification was performed using LVEF and left atrial volume index (LAVI) as surrogate markers of LV and LA reverse remodelling, respectively. Analogous to the strain-based approach, patients were classified into four groups according to the direction of change in LVEF and LAVI between baseline and one-year follow-up, and clinical outcomes were compared across these volume-based recovery phenotypes.

To identify independent predictors of LV recovery (ΔLVGLS >0), LA recovery (ΔLASr >0), and concordant recovery (ΔLVGLS >0 and ΔLASr >0), univariable logistic regression analyses were first performed for each recovery phenotype using baseline clinical and echocardiographic parameters. Variables achieving statistical significance (p<0.05) on univariable analysis were subsequently entered into multivariable logistic regression models, with results expressed as adjusted odds ratios (OR).

As an exploratory analysis, LV compliance was additionally assessed as a candidate predictor in the logistic regression models. Left ventricular end-diastolic pressure (LVEDP) was estimated as 11.96 + 0.596 × E/e’,^14^ and the individual end-diastolic pressure-volume relationship was parameterized from the estimated LVEDP and left ventricular end-diastolic volume (LVEDV) using the Klotz single-beat method.^15,16^ LV compliance was quantified as the LVEDV at a LVEDP of 30 mmHg (V30mmHg).

All statistical analyses were performed using Python (version 3.12.3; Python Software Foundation). Two-sided p values <0.05 were considered statistically significant.

## 3. RESULTS

### 3.1 Study Population and Baseline Characteristics

A total of 1,182 patients were included in the final analysis (**Figure 1**). Based on the recovery patterns in LVGLS and LASr at one-year follow-up, patients were classified into Group A (concordant recovery, n=677, 57.4%), Group B (discordant atrial non-recovery, n=132, 11.2%), Group C (discordant ventricular non-recovery, n=184, 15.6%), and Group D (concordant non-recovery, n=189, 16.0%).

Baseline characteristics across the four groups are summarized in **Table 1**. Groups B and D were older (median age 69.5 and 69.9 years, respectively) compared with Groups A and C (64.6 and 65.9 years). The prevalence of hypertension and diabetes mellitus was highest in Group B (43.9% and 36.4%, respectively), while coronary artery disease was most prevalent in Groups B and C (41.7% and 44.0%, respectively). The prevalence of atrial fibrillation was similar across the four groups (22.9%, 22.7%, 22.3%, and 26.5% in Groups A, B, C, and D, respectively). NT-proBNP levels were notably lower in Group B (845 pg/mL [IQR 476–2,673]) compared with the other groups. Concomitant medication use did not differ significantly across groups, with the exception of diuretic use, which was highest in Group A (68.5%, p=0.004).

**Table 1.**
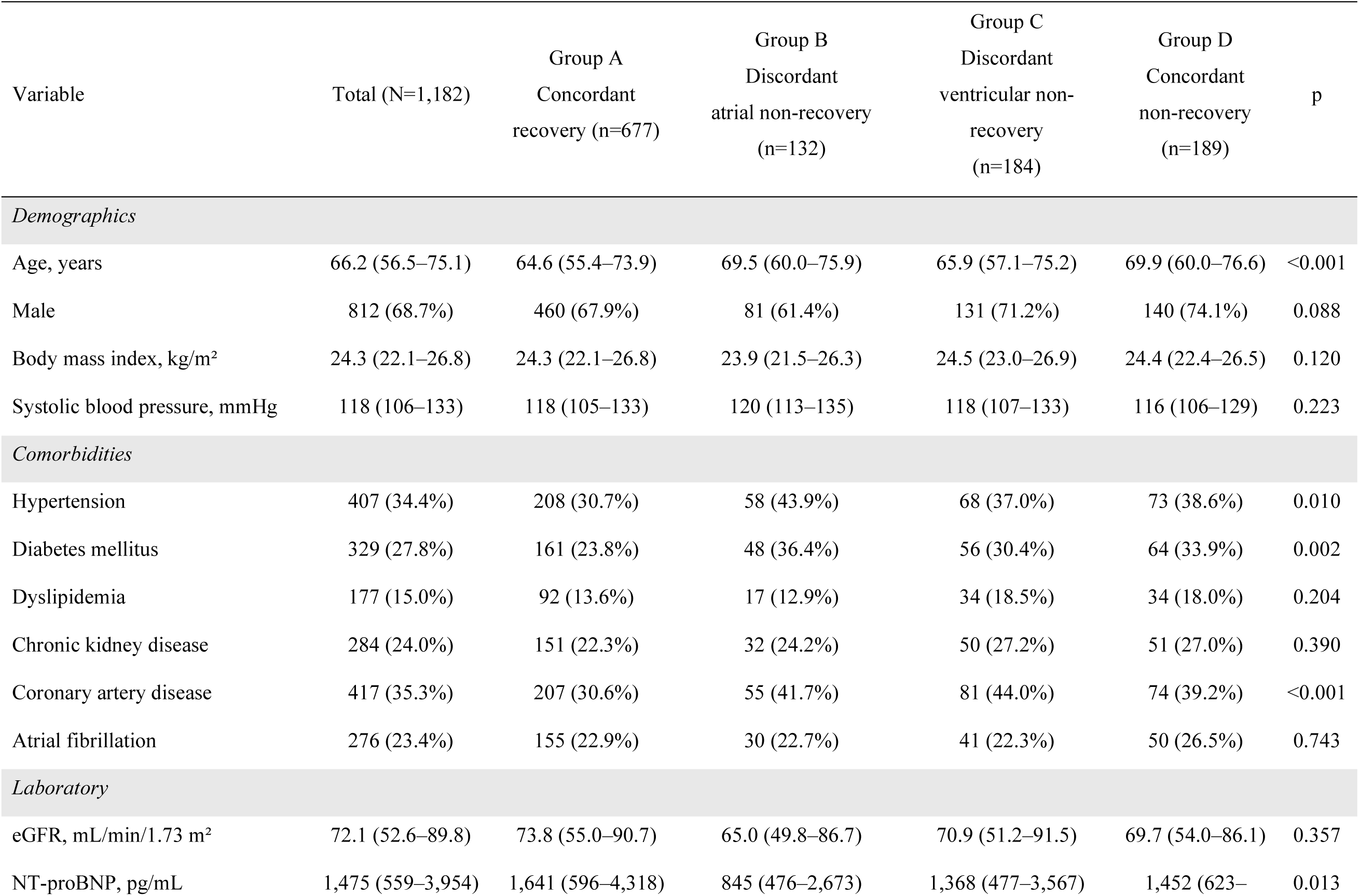
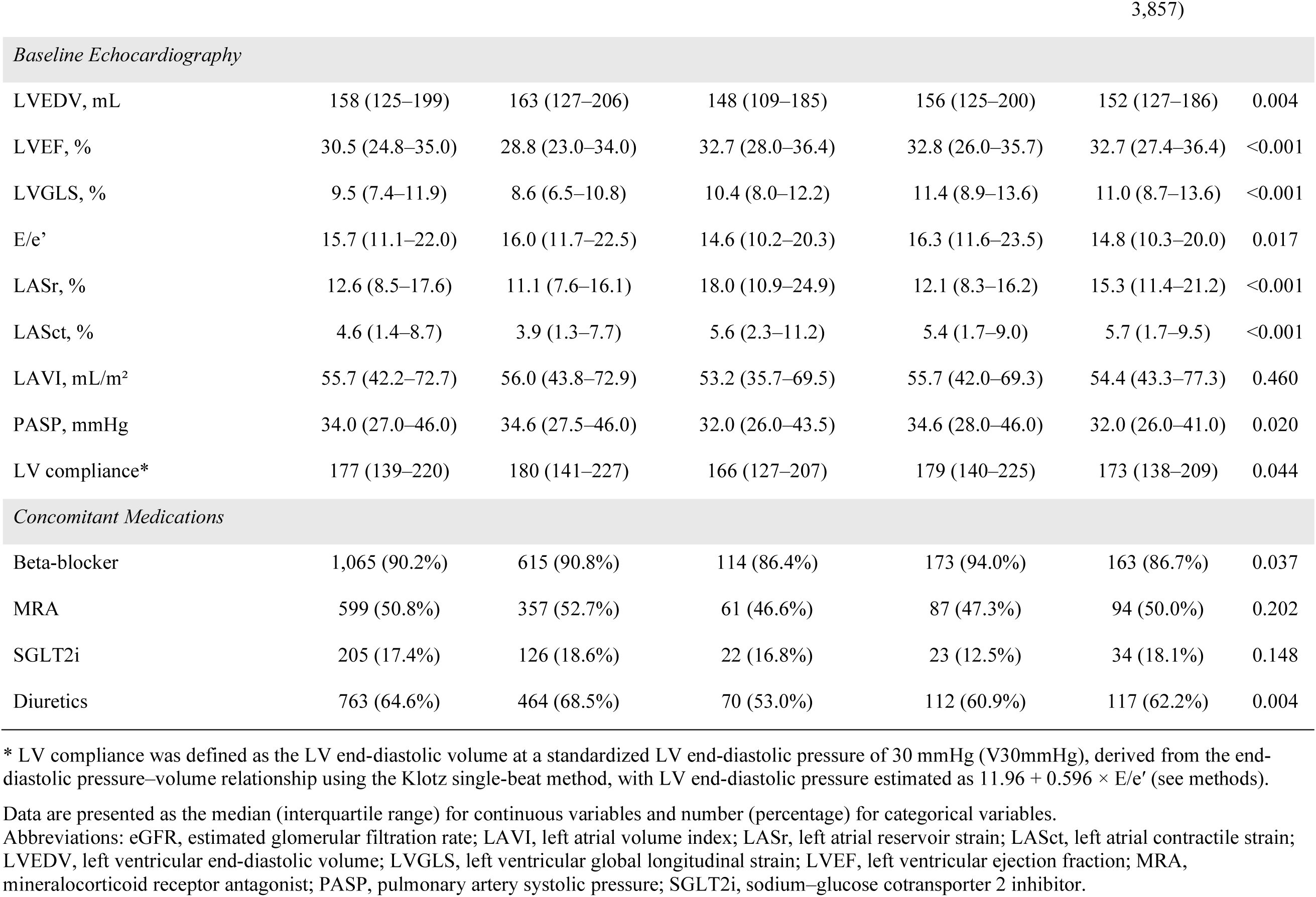
Baseline Characteristics by Strain Recovery Group.

Baseline echocardiographic parameters differed significantly across groups. Group A exhibited the most advanced myocardial dysfunction at baseline, with the lowest LVEF (28.8% [23.0–34.0]), LVGLS (8.6% [6.5–10.8]), and LASr (11.1% [7.6–16.1]), and the largest LVEDV (163 mL [127–206]). Group D, the concordant non-recovery group, demonstrated higher baseline LVEF (32.7% [27.4–36.4]) and LVGLS (11.0% [8.7–13.6]) compared with Group A. Group B had the highest baseline LASr among all groups (18.0% [10.9–24.9]), though baseline LVGLS was comparable to Groups C and D.

### 3.2 Serial Changes in LVGLS and LASr

Serial changes in LVGLS and LASr differed markedly across the four strain recovery groups (**Figure 2**). LVGLS improved significantly from baseline to one year in Groups A and B, whereas it declined significantly in Groups C and D (**Figure 2A**). The magnitude of LVGLS improvement was greater in Group A than in Group B, and conversely, the magnitude of LVGLS decline was more pronounced in Group D than in Group C (**Figure 2B**). Regarding LASr, significant improvement from baseline to one year was observed in Groups A and C, whereas LASr declined in Groups B and D (**Figure 2C**). The extent of LASr recovery was attenuated in Group C, the discordant group with LVGLS non-recovery, compared with Group A. Notably, despite having the highest baseline LASr among all groups, Group B exhibited a degree of LASr deterioration comparable to that of Group D (**Figure 2D**). The joint distribution of LVGLS and LASr showed largely overlapping profiles at baseline (**Figure 2E**). At one-year follow-up, however, the groups became more distinctly separated, with Group B clustering in a region of improved LVGLS but persistently low LASr—a pattern visually distinct from the concordant changes observed in Groups A and D (**Figure 2F**).

**Figure 2.**
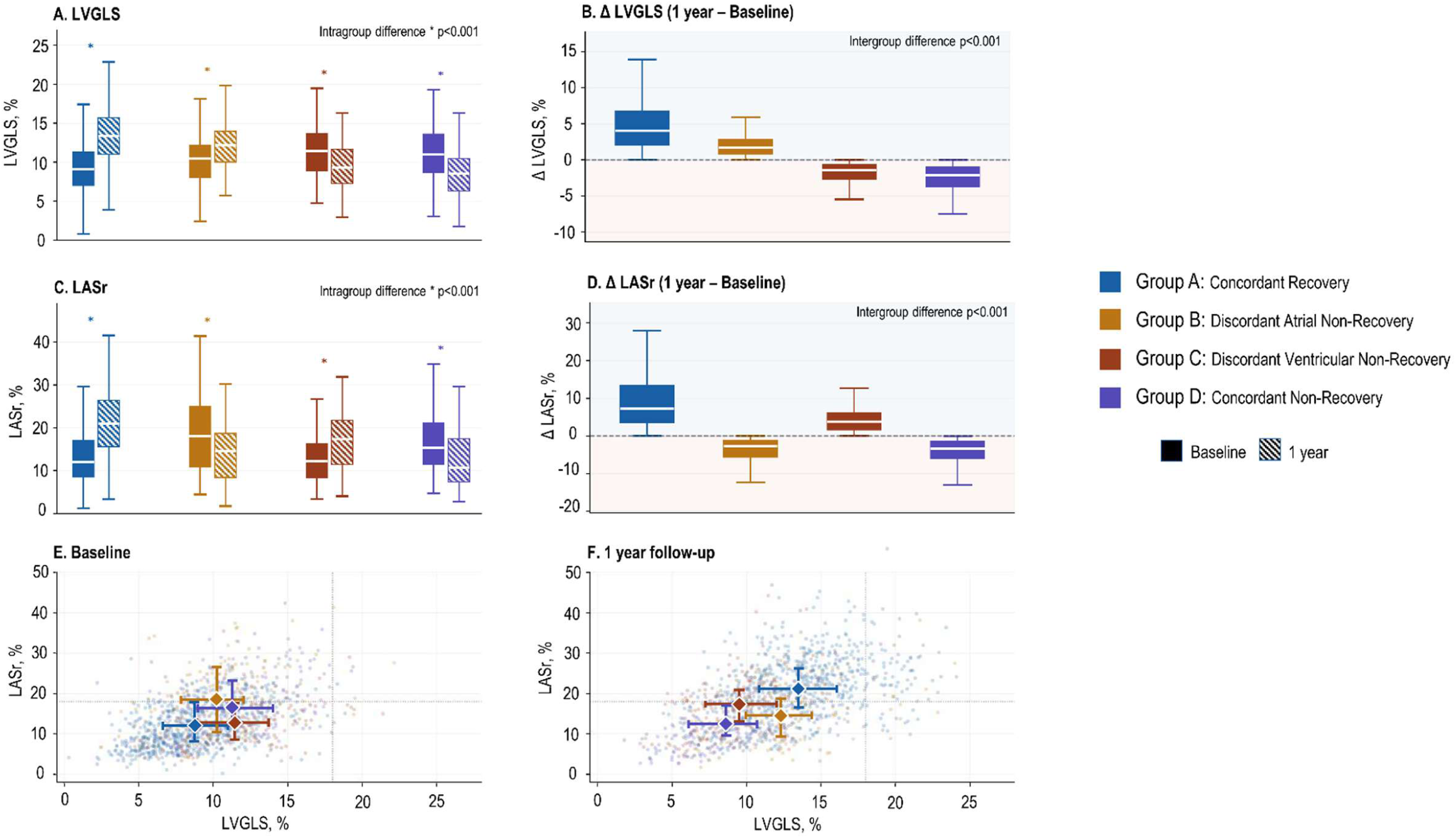
Serial Changes in Left Ventricular and Left Atrial Strain According to Strain Recovery Pattern. (A) Serial trajectories of LVGLS from baseline to one-year follow-up by strain recovery group, presented as median with interquartile range. (B) Between-group differences in ΔLVGLS. (C) Serial trajectories of LASr from baseline to one-year follow-up. (D) Between-group differences in ΔLASr. (E) Joint distribution of LVGLS and LASr at baseline. (F) Joint distribution of LVGLS and LASr at one-year follow-up. Diamond markers indicate group medians. LASr, left atrial reservoir strain; LVGLS, left ventricular global longitudinal strain.

### 3.3 Clinical Outcomes

Over a median post-landmark follow-up of 26.0 months (IQR 15.2–40.3), 100 all-cause deaths, 56 cardiovascular deaths, and 114 HF hospitalizations occurred. When LVGLS and LASr changes were analysed as continuous variables (**Table 2**), each 5% decrease in LVGLS was significantly associated with cardiovascular mortality (adjusted hazard ratio [aHR] 2.00, 95% CI 1.29–3.10), but not with all-cause mortality (aHR 1.28, 95% CI 0.93–1.78). In contrast, each 5% decrease in LASr was independently associated with both all-cause mortality (aHR 1.52, 95% CI 1.23–1.89) and cardiovascular mortality (aHR 1.87, 95% CI 1.39–2.52), highlighting the incremental prognostic contribution of LA functional deterioration beyond that of LV strain.

**Table 2.** Association Between Serial Strain Changes and Clinical Outcomes.

| Strain Changes | Adjusted HR (95% CI) | p-value |
| --- | --- | --- |
| Worsening LVGLS, per 5% decrease |  |  |
| All-cause death | 1.28 (0.93–1.78) | 0.129 |
| Cardiovascular death | 2.00 (1.29–3.10) | 0.002 |
| Heart failure hospitalization | 1.32 (0.98–1.77) | 0.066 |
| Worsening LASr, per 5% decrease |  |  |
| All-cause death | 1.52 (1.23–1.89) | <0.001 |
| Cardiovascular death | 1.87 (1.39–2.52) | <0.001 |
| Heart failure hospitalization | 1.17 (0.98–1.40) | 0.075 |

Abbreviations: LASr, left atrial reservoir strain; LVGLS, left ventricular global longitudinal strain.

Kaplan-Meier curves for clinical outcomes showed significant differences across the four strain recovery groups (**Figure 3**). For both all-cause and cardiovascular mortality, Group A (concordant LASr and LVGLS recovery) had the lowest event rates, while Group D (concordant LASr and LVGLS non-recovery) had the highest. Of note, Group B (discordant LASr non-recovery) demonstrated markedly higher mortality rates compared with Group A, despite having achieved LV functional improvement. In addition, the mortality rates in Group B were higher than those in Group C (discordant LVGLS non-recovery). For HF hospitalization, Group A again showed the lowest event rate, whereas Group C showed a higher event rate comparable to that of Group D.

**Figure 3.**
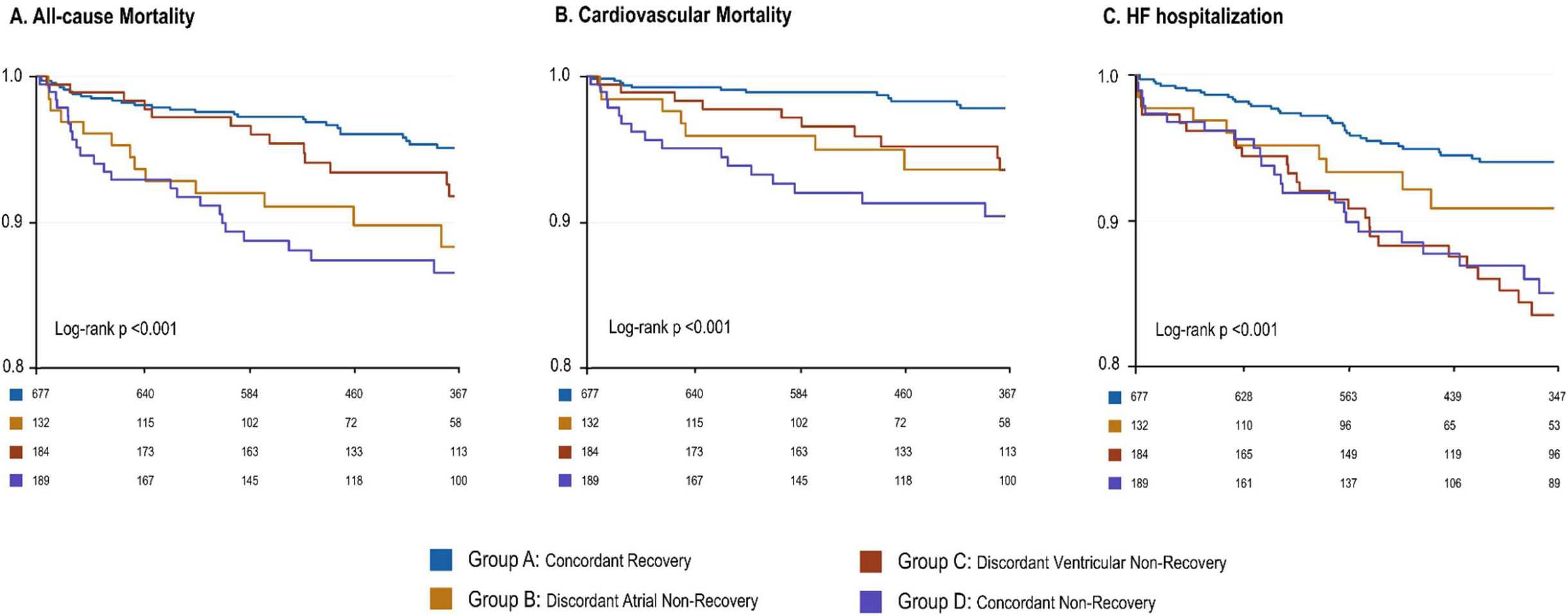
Clinical Outcomes by Strain Recovery Group. Kaplan-Meier curves for (A) all-cause mortality, (B) cardiovascular mortality, and (C) HF hospitalization according to strain recovery group. Index date was the one-year follow-up echocardiography. HF, heart failure.

On multivariable Cox regression analysis (**Figure 4**), Group B was associated with significantly higher risks of all-cause mortality (aHR 3.53, 95% CI 1.60–7.79) and cardiovascular mortality (aHR 5.68, 95% CI 1.91–16.92) compared with Group A. These risks were comparable to those observed in Group D, which showed similarly elevated all-cause mortality (aHR 3.35, 95% CI 1.81–6.18) and cardiovascular mortality (aHR 6.00, 95% CI 2.60–13.87). Group C demonstrated a significantly higher risk of HF hospitalization (aHR 2.25, 95% CI 1.31–3.86) and elevated cardiovascular mortality (aHR 2.53, 95% CI 1.00–6.39) compared with Group A.

**Figure 4.**
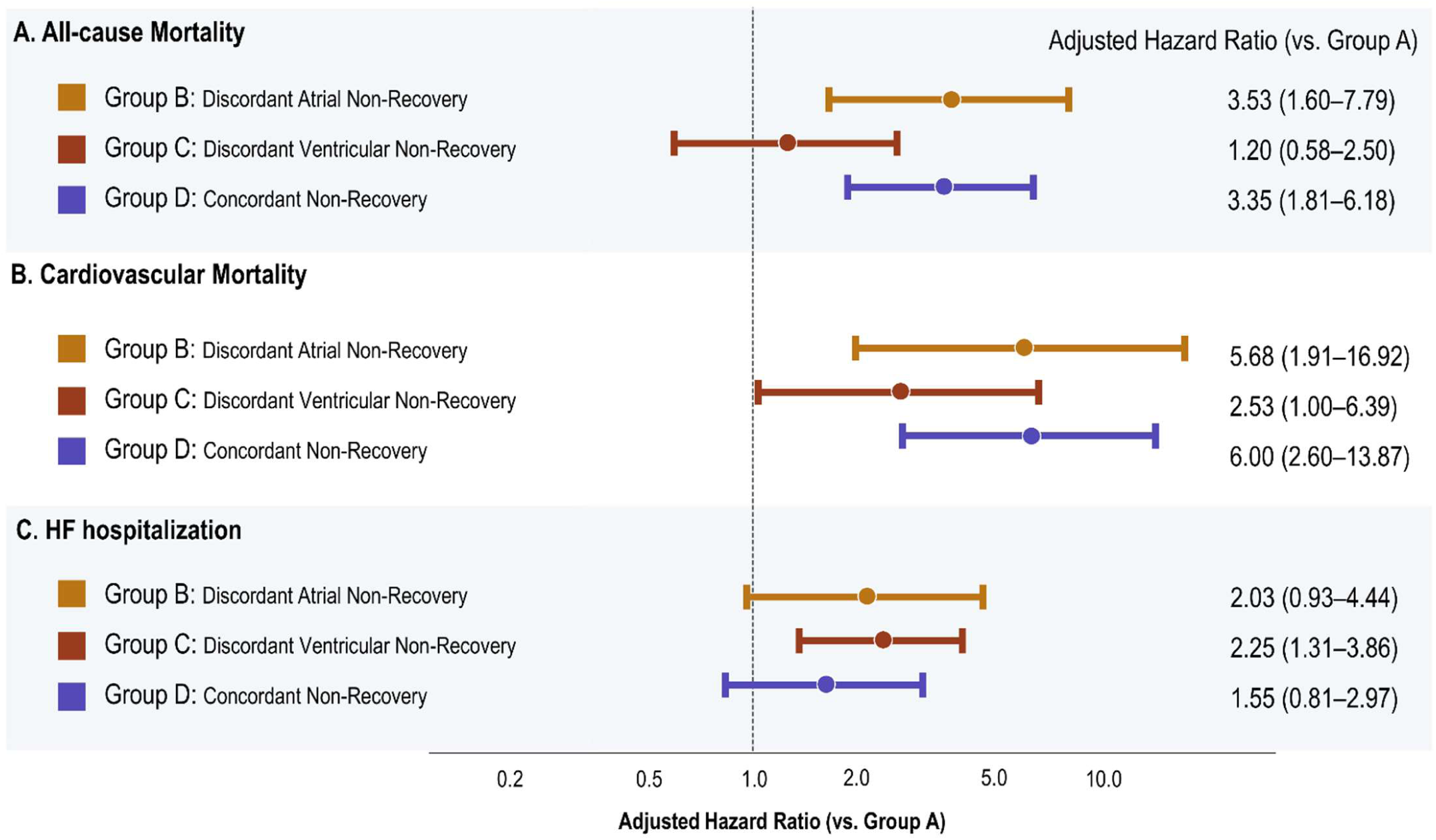
Adjusted Hazard Ratios for Clinical Outcomes by Strain Recovery Group. Forest plots showing unadjusted and adjusted hazard ratios for (A) all-cause mortality, (B) cardiovascular mortality, and (C) HF hospitalization according to strain recovery group, with Group A (concordant recovery) as the reference. Multivariable models were adjusted for age, sex, baseline LVEF, NT-proBNP, ischemic cardiomyopathy, chronic kidney disease, and concomitant SGLT2 inhibitor use. Data are presented as hazard ratio (95% confidence interval). HF, heart failure.

AF prevalence at the 1-year landmark timepoint did not differ significantly across the four groups (Group A 29.0%, Group B 30.8%, Group C 28.3%, Group D 31.6%; p=0.866; **Table S1**). In a sensitivity analysis focused on patients without baseline AF (n=905), the adverse prognostic impact of Group B was preserved, with all-cause mortality (aHR 3.33, 95% CI 1.28–8.66) and cardiovascular mortality (aHR 7.06, 95% CI 1.84–27.17) remaining significantly elevated compared with Group A. The elevated mortality risk in Group D was similarly maintained, whereas Group C did not show significant mortality associations in this subgroup (**Table S2**).

### 3.4 Hemodynamic Trajectories in Discordant Atrial Non-Recovery

To further characterize the excess mortality risk observed in Group B despite apparent LV recovery, hemodynamic trajectories between Groups A and B were compared (**Figure S1**). Despite comparable LVEF trajectories at one year, Group B showed less favourable improvements in LA remodelling and hemodynamic parameters, including LAVI, E/e’, PASP, and NT-proBNP, compared with Group A. In an exploratory analysis of LV compliance based on LVEDV and estimated LVEDP, the magnitude of LV reverse remodelling, as reflected by the reduction in LVEDV and LVEDP was attenuated in Group B than in Group A (intergroup difference p<0.001; **Figure S2**). These findings suggest that persistent LA dysfunction in Group B is accompanied by ongoing hemodynamic burden that may underlie the worse clinical outcomes.

Serial assessment of LASct across all four groups further corroborated this pattern (**Figure S3**). While Group A demonstrated substantial improvement in LASct at 1 year (Δ+3.50%, p<0.001), Group B showed no meaningful recovery in LASct (Δ+0.00%), and Group D exhibited further deterioration (Δ-0.56%, p<0.001). The between-group difference in ΔLASct was significant (intergroup difference p<0.001).

### 3.5 Subgroup Analysis by Baseline Strain Level

The prognostic significance of strain recovery patterns was further examined according to baseline levels of LVGLS and LASr (**Figure S4**). When stratified by baseline LVGLS, the adverse prognostic impact of discordant atrial non-recovery remained consistent across subgroups, with Group B demonstrating higher mortality risk compared with Group A. Similarly, when stratified by baseline LASr, the excess mortality risk in Group B showed consistent trends across subgroups with high and low LASr at baseline.

### 3.6 Comparison with Volume-Based Recovery Classification

Under volume-based recovery classification using LVEF and LAVI, the concordant non-recovery group (Group D; decreased LVEF and increased LAVI) remained associated with significantly elevated mortality risk (all-cause mortality aHR 4.26, 95% CI 2.01–9.00) (**Figure S5**). However, the discordant atrial non-recovery group (Group B; improved LVEF but increased LAVI) did not demonstrate significantly elevated all-cause mortality (aHR 1.34, 95% CI 0.65–2.75) or cardiovascular mortality risk (aHR 1.37, 95% CI 0.47–3.95). This contrasted with the markedly elevated mortality risk observed in Group B under strain-based classification.

### 3.7 Predictors of Strain Recovery

Independent predictors of LV recovery, LA recovery, and concordant recovery are shown in **Figure 5**. For LV recovery, younger age, absence of CAD, and lower baseline LVGLS emerged as independent predictors on multivariable analysis. For LA recovery, lower baseline LASr was the strongest predictor (OR 1.97 per 5% decrease, 95% CI 1.68–2.30), with beta-blocker use, absence of diabetes mellitus, and younger age also independently associated. Higher baseline LV compliance was additionally associated on univariate analysis (OR 1.05 per 20 mL increase, 95% CI 1.01–1.10), suggesting that greater LV compliance at baseline may facilitate LA reverse remodelling, although this association did not persist after multivariable adjustment. Concordant recovery shared younger age as a common predictor with both LV and LA recovery, with lower baseline LASr serving as an additional independent determinant. As with LA recovery, higher baseline LV compliance showed univariate association with concordant recovery (OR 1.05, 95% CI 1.01–1.09) without reaching multivariable significance.

**Figure 5.**
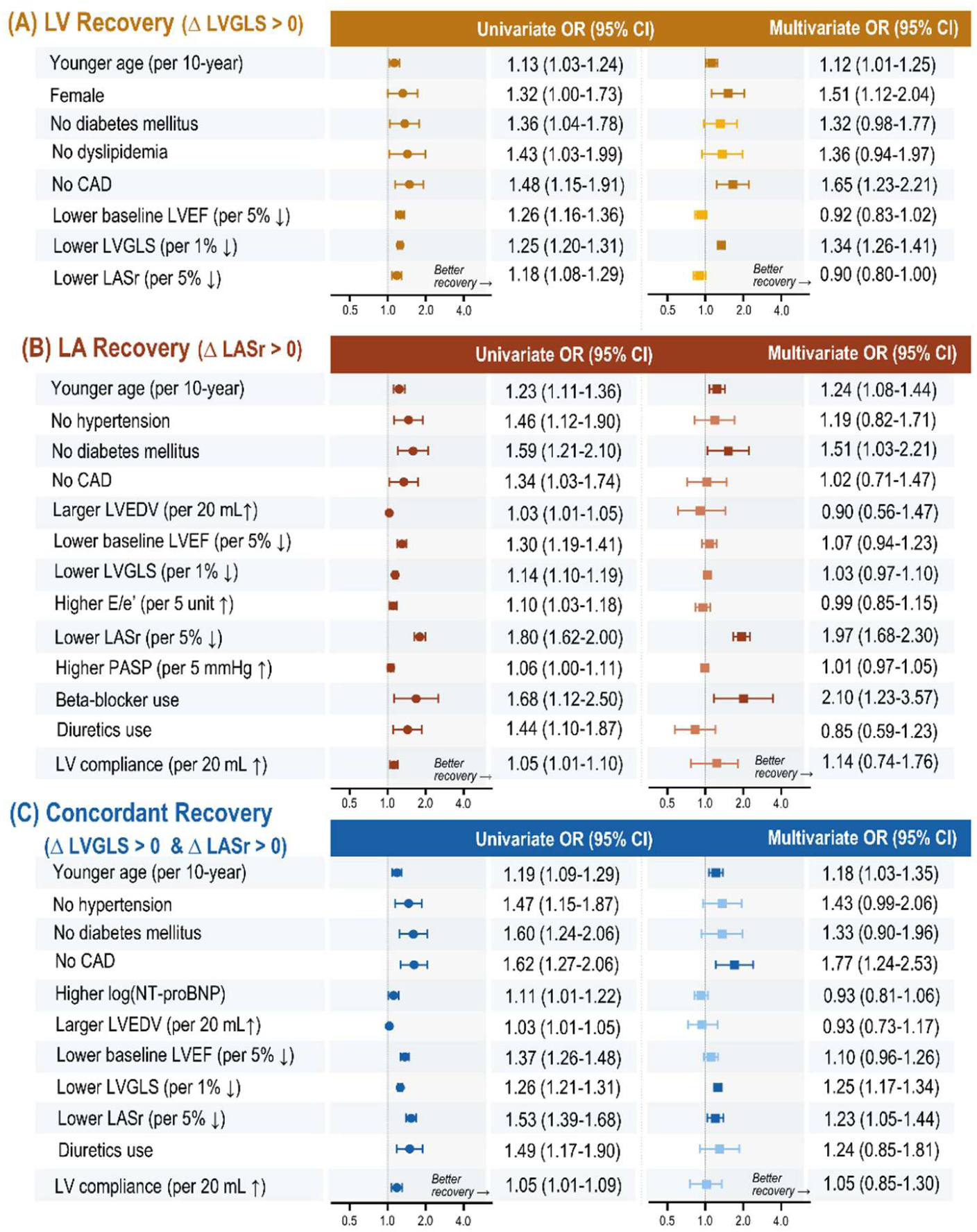
Predictors of LV, LA, and Concordant Recovery. Forest plots showing univariable and multivariable odds ratios for independent predictors of (A) LV recovery (ΔLVGLS >0), (B) LA recovery (ΔLASr >0), and (C) concordant recovery (ΔLVGLS >0 and ΔLASr >0). LV compliance was derived as LV volume at a left ventricular end-diastolic pressure of 30 mmHg, estimated from the end-diastolic pressure-volume relationship. Variables achieving p<0.05 on univariable analysis were entered into multivariable models. CAD, coronary artery disease; CI, confidence interval; E/e’, ratio of early transmitral flow velocity to early diastolic annular velocity; LASr, left atrial reservoir strain; LVEDV, left ventricular end-diastolic volume; LVEF, left ventricular ejection fraction; LVGLS, left ventricular global longitudinal strain; NT-proBNP, N-terminal pro–B-type natriuretic peptide; OR, odds ratio; PASP, pulmonary artery systolic pressure.

## 4. DISCUSSION

In this multicentre real-world registry of HFrEF patients treated with ARNI-based therapy, we characterized four distinct strain recovery phenotypes based on concurrent changes in LVGLS and LASr at one-year follow-up. The principal finding of this study is that discordant atrial non-recovery—defined by the absence of LA recovery despite LV functional improvement—was associated with mortality risk comparable to concordant non-recovery. This pattern of “deceptive recovery” was accompanied by persistently unfavourable hemodynamic trajectories and was consistently observed regardless of baseline strain severity. LV recovery was independently predicted by younger age, absence of CAD, and lower baseline LVGLS, reflecting the role of residual ventricular contractile reserve at treatment initiation. LA recovery, in contrast, was most strongly predicted by lower baseline LASr, along with beta-blocker use and absence of diabetes mellitus, suggesting that the capacity for atrial functional recovery is affected by distinct clinical and mechanical determinants than those from LV. These findings suggest that persistent LA dysfunction despite LV functional improvement identifies a high-risk phenotype, and that concurrent assessment of LV and LA strain may provide incremental prognostic value beyond LV-centric metrics alone in HFrEF.

The observation that discordant atrial non-recovery, despite apparent improvement in LV function, confers a mortality risk higher than that of discordant ventricular non-recovery and even comparable to that of concordant non-recovery challenges the conventional paradigm of treatment response assessment in HFrEF. Prior studies evaluating reverse remodelling under ARNI-based therapy have predominantly focused on LV-centric parameters, demonstrating that improvement in LVEF and LVGLS is associated with favourable clinical outcomes.^2,7,17^ However, these analyses have been largely LV-centric, and the concurrent trajectory of LA function has received limited attention. The present findings suggest that while LV functional improvement is an important marker of treatment response, concurrent LA recovery may have substantial contribution in determining long-term prognosis. When strain changes were analysed as continuous variables, both worsening LVGLS and worsening LASr were independently associated with cardiovascular mortality, whereas LASr deterioration carried additional prognostic weight for all-cause mortality. These findings highlight the independent and complementary prognostic effect of LA recovery beyond LV improvement in HFrEF. The mechanistic basis underlying persistent LA dysfunction despite LV functional improvement in Group B warrants consideration. A key observation in the present study is that Group B had the highest baseline LASr among all groups, yet exhibited a degree of LASr deterioration at one year comparable to that of Group D. This finding suggests that the failure of LA recovery in Group B is not attributable to pre-existing severe atrial dysfunction, but rather reflects an underlying atrial myopathy that is less amenable to reverse remodelling divergent to LV recovery.

LA dysfunction in HFrEF is driven by a complex interplay of chronic pressure overload, atrial fibrosis, and neurohormonal activation, which may progress independently of LV systolic function.^4,18^ The clinical profile of Group B further supports an established atrial cardiomyopathy (AtCM) substrate underlying this hemodynamic pattern. Compared with Group A, Group B patients were older, had higher burdens of diabetes mellitus, hypertension, and renal dysfunction, all recognized risk factors for atrial structural remodelling and AtCM progression.^19^ Among all four groups, Group B carried the highest prevalence of both hypertension and diabetes mellitus, exceeding even Group D, the group with the most adverse overall recovery phenotype. The independent predictors of LA recovery identified in **Figure 5**, namely younger age, absence of diabetes mellitus, and beta-blocker use, are precisely the characteristics that distinguished Group B from Group A, suggesting that LA non-recovery reflects a pre-existing atrial substrate that exceeds the reversibility achievable through ventricular unloading alone. We propose that this atrial substrate, driven by the accumulation of recognized AtCM risk factors, impairs the improvement of diastolic hemodynamic parameters (E/e’, LAVI, PASP) even when LV systolic function recovers, and it is this persistent hemodynamic burden that is ultimately captured as LA strain non-recovery at the 1-year assessment. Although direct quantification of LA fibrosis by cardiac MRI or invasive measurement of LA pressure was not performed in the present registry, LASr provides a sensitive and clinically applicable surrogate of LA mechanical dysfunction, and its use in guideline-recommended echocardiographic assessment reflects its established utility for detecting subclinical atrial myopathy.^7,13,20^

The hemodynamic trajectory data further support this interpretation: despite comparable LVEF improvement, Group B demonstrated persistently elevated filling pressures and less favourable LA remodelling, as reflected by E/e’, LAVI, PASP, and NT-proBNP, compared with Group A. Additionally, the attenuated reduction in LVEDP and LVEDV in Group B, indicative of less favourable reverse remodelling, provides further evidence that residual diastolic burden persists despite apparent LV systolic recovery. As previously described in patients with HFrEF receiving ARNI therapy,^16^ Group A exhibited a substantially greater leftward and downward shift of the LV pressure–volume relationship, whereas Group B showed an attenuated reverse remodelling trajectory. Together, these hemodynamic findings indicate that persistent elevation of LV filling pressure, even in the presence of partial LV systolic recovery, is mechanistically important. Chronically elevated filling pressures perpetuate increased atrial afterload and impaired atrial compliance, thereby preventing LA reservoir function recovery and promoting the progressive atrial structural remodelling that constitutes the substrate of AtCM. This persistent hemodynamic burden, even in the context of apparent ventricular recovery, may represent the structural substrate for the excess mortality risk observed in Group B. Consistent with this interpretation, serial assessment of LASct demonstrated that Group B showed no meaningful improvement at 1 year, in contrast to Group A. These findings indicate that persistent LA dysfunction in Group B encompasses both reservoir and contractile phases of LA mechanical function, providing additional mechanistic evidence for an established AtCM substrate that is insufficiently reversible through ventricular unloading alone. The persistent LA dysfunction characterizing Groups B and D in the present study may be conceptualized within the framework of AtCM, as defined by the 2024 EHRA consensus statement, which encompasses structural, functional, and electrophysiological atrial changes with adverse clinical implications.^19^ In the EAST-AFNET 4 trial, advanced AtCM was independently associated with higher rates of cardiovascular events, consistent with the excess mortality risk observed in patients without LA recovery in our cohort.^21^ Together, these findings suggest that LASr trajectory may serve as a functional marker of AtCM severity and progression in HFrEF patients. Additionally, our findings may facilitate further studies characterizing LA phenotypes and the clinical relevance of AtCM using LA strain measurements and complementary indices, such as the LA stiffness index, which integrates LV filling pressure and LA mechanical function.

In the sensitivity analysis using volume-based recovery classification, Group B—defined by improved LVEF but increased LAVI—did not demonstrate significantly elevated mortality risk, in contrast to the markedly elevated risk observed under strain-based classification. This discrepancy may reflect an inherent limitation of volumetric parameters in capturing LA myopathy that develops independently of LV remodelling. Unlike LAVI, which reflects structural enlargement, LASr provides a more sensitive assessment of LA functional reserve and may therefore identify residual atrial dysfunction.^20^

The distinct predictor profiles of LV and LA recovery provide further mechanistic insight into their divergent trajectories. For both LV and LA recovery, lower baseline strain values—lower LVGLS and lower LASr, respectively—were independent predictors, suggesting that patients with greater baseline impairment retain more potential for functional improvement with ARNI-based therapy, a pattern consistent with prior reverse remodelling studies.^9,17^ Beyond this shared baseline effect, however, the two recovery phenotypes were governed by distinct clinical determinants. LV recovery was additionally predicted by younger age and absence of CAD, suggesting that the extent of irreversible myocardial injury and fibrotic substrate limits ventricular plasticity.^22^ In contrast, LA recovery was independently associated with beta-blocker use and absence of diabetes mellitus, in addition to lower baseline LASr. The association of beta-blocker use with LA recovery may reflect the beneficial effects of heart rate reduction and sympathetic inhibition on atrial remodelling, which modulate LA mechanics independently of LV functional improvement.^23^ The absence of diabetes mellitus as a predictor of LA recovery is consistent with evidence that metabolic comorbidities impair atrial myocardial plasticity and attenuate the potential for functional recovery.^23,24^ These findings suggest that LV and LA recovery are governed by partially distinct pathophysiological pathways, highlighting the clinical relevance of targeting atrial functional recovery beyond LV-centric treatment strategies.

The findings of the present study carry several clinical implications. First, the identification of discordant atrial non-recovery as a high-risk phenotype suggests that serial assessment of LASr alongside LVGLS may enhance risk stratification in HFrEF patients. Current monitoring strategies predominantly rely on LVEF and NT-proBNP as treatment response markers^25^; however, these parameters may fail to identify patients who achieve apparent LV recovery yet harbour persistent LA myopathy with adverse prognostic implications. Second, the ability of LASr to detect residual hemodynamic burden independent of volumetric parameters supports its role as a complementary imaging biomarker in the follow-up of HFrEF. Incorporating LASr into echocardiographic follow-up may provide actionable information for identifying patients who may benefit from more intensive monitoring or further optimization of medical therapy.^17^ Third, the distinct predictor profiles of LV and LA recovery suggest that a subset of patients—particularly those who are older or have diabetes mellitus—may be at risk for discordant atrial non-recovery despite favourable ventricular response, and may warrant closer surveillance.

Notwithstanding these clinical implications, certain limitations of the present study should be acknowledged. First, the retrospective nature of the analysis introduces inherent selection bias, as patients without available serial TTE data were excluded, which may limit the generalizability of the findings. Second, all data were obtained from two tertiary referral centres in Korea, and the extent to which these findings apply to broader and more diverse HFrEF populations remains to be established. Third, the strain recovery classification was based on the direction of change in LVGLS and LASr at a single one-year timepoint, and does not capture the dynamic trajectories that may occur over longer follow-up periods. Fourth, the mechanisms underlying discordant atrial non-recovery, including the extent of atrial fibrosis and neurohormonal activation, were not directly assessed in the present study, and further investigations incorporating cardiac magnetic resonance imaging or invasive hemodynamic measurements would be warranted to elucidate the pathophysiological basis of this phenotype. Furthermore, formal staging of AtCM according to the 2024 EHRA consensus criteria was not feasible in the present cohort, as PR interval data were not systematically collected in the registry. Finally, AF pattern (paroxysmal, persistent, or permanent) was not systematically recorded in the registry, precluding an analysis of its differential influence on LA functional recovery and AtCM progression. Future studies incorporating structured AF pattern classification would allow a more nuanced examination of the interplay between AF burden and discordant atrial non-recovery.

## 5. CONCLUSION

In HFrEF patients treated with ARNI-based therapy, discordant atrial non-recovery—characterized by persistent LA dysfunction despite LV functional improvement on strain echocardiography—was associated with mortality risk comparable to concordant non-recovery. This pattern of deceptive recovery was accompanied by persistently unfavourable hemodynamic trajectories and was not captured by conventional volumetric parameters. Concurrent assessment of LV and LA strain provides incremental prognostic information beyond LV-centric metrics, and thereby enhances the identification of high-risk phenotypes in HFrEF patients.

## Data Availability

The data underlying this study cannot be made publicly available due to ethical restrictions set by the IRB of the study institution; i.e., public availability would compromise patient confidentiality and participant privacy. Please contact the corresponding author to request the minimal anonymized dataset.

